# Improving fidelity of self-administered pulse oximetry and remote patient monitoring implementation in Honduras during the COVID-19 pandemic

**DOI:** 10.1101/2025.07.03.25330771

**Authors:** Kathryn W. Roberts, Berta Alvarez, Michael de St. Aubin, Omar Diaz, Salomé Garnier, Rachel See, Saul Cruz, Lorenzo Pavon, Anthony D. So, Ligia Paina, Tara Kirk Sell, Shiony Midence, Angela Ochoa, Homer Mejía Santos, Jonatán Ochoa, Sogeiry Solis, Devan Dumas, Margaret Baldwin, C. Daniel Schnorr, Alcides Martinez, Avi Hakim, Eric Nilles

## Abstract

During the COVID-19 pandemic, Honduras’ Secretariat of Health was eager to understand whether self-administered pulse oximetry could improve clinical outcomes of high risk COVID-19 patients. While these interventions show promise for expanding healthcare access in lower- and middle-income countries, evidence about contextual implementation fidelity and effectiveness remains limited.

We conducted a retrospective fidelity of implementation analysis of phone-based patient monitoring with or without self-administered pulse oximetry in Tegucigalpa and Comayagüela, Honduras. Using Carroll’s Conceptual Framework for Implementation Fidelity, we analyzed adherence (content, coverage, frequency, duration), delivery quality, participant responsiveness, and implementation strategies through trial data, acceptability assessments, and implementation documentation.

The intervention achieved high fidelity: 97.9% coverage (1,821/1,860 eligible patients), 97.7% daily monitoring call completion calls, and 97.8% pulse oximeter utilization. Implementation strategies including culturally-adapted education materials and communication styles, and multi-disciplinary team engagement. Participant satisfaction was 99.7%, with minimal withdrawal (0.1%) and loss to follow-up (2.0%). However, only 28% of participants sought additional care when referred for warning signs. Unexpected benefits included reported positive impacts on participants’ mental health during isolation.

This analysis demonstrates that remote patient monitoring and self-administered pulse oximetry implementation can achieve high fidelity in a lower-middle-income setting, including during a health emergency and where such interventions are novel. This challenges concerns about technology access and adoption in resource-limited environments. Future research should focus on hybrid effectiveness-implementation trials and strategies to improve referral adherence. These findings suggest that expanding implementation of these approaches may enhance healthcare access during emergencies and routine circumstances.

## Introduction

In 2020, in response to the COVID-19 pandemic, Honduras’ Secretariat of Health (SESAL) established dedicated triage centers to streamline care and manage the increasing patient load.^1^ To further expand healthcare access, SESAL embraced home-based care guidelines developed by the World Health Organization (WHO), which provided guidance for remote care delivery for high-risk, non-hospitalized patients.^2^ In April 2021, the WHO added self-administered pulse oximetry to previous guidelines, advising use to detect silent hypoxia (clinical deterioration without shortness of breath or other danger signs).^3^ SESAL adapted the intervention to align with the context, technology penetration, telehealth experience, ability to self-isolate, and availability of household members to provide support.^2,4^ The U.S. Centers for Disease Control (CDC) supported remote patient monitoring and self-administered pulse oximetry as part of its emergency pandemic response. Remote patient monitoring and self-administered pulse oximetry have been evaluated in previous COVID-19 and other, but additional evidence was needed, particularly in lower- and middle-income country (LMIC) contexts. In response, SESAL and partners implemented a randomized trial and sub-studies to establish clinical effectiveness, implementation fidelity, and acceptability.^5–11^

Honduras has a rich and intricate social and political landscape, and local context was critical to intervention and implementation design. As a lower-middle-income country, it faces challenges such as governmental issues, organized crime, high rates of violence, and struggles within its healthcare system.^12–17^ This study took place in Tegucigalpa, the capital of Honduras, and its sister city, Comayagüela, which had a combined population of 1.3 million in 2023, with 22,000 residents per square mile, many of whom live in densely crowded informal settlements, facilitating SARS-CoV-2 transmission.[4],[5] Honduras is the most economically unequal country in Latin America and 45.6% of its economy is informal, which can influence availability of space or financial buffer to isolate, socially distance, or afford preventive measures. [6],[7] Ninety percent of Hondurans access healthcare for free through the public sector.^22,23^ However, this healthcare system experiences challenges aligning care availability and quality with patient needs, due to gaps in funding, staffing, and commodities.[7],[8] Remote patient monitoring and self-administered pulse oximetry were not part of usual care in Honduras prior to study implementation. These interventions provided a means to deliver support to non-hospitalized COVID-19 patients at high risk of adverse outcomes and the opportunity to examine the utility and feasibility of these interventions in the Honduran and health emergency contexts, as a complement to existing in-person and community-based healthcare provision modalities.

In collaboration with SESAL, the study management team tailored strategies to enhance implementation, delivery quality, and participant responsiveness to optimize fidelity – the degree to which a program is delivered as intended. Implementation fidelity can be challenging in contexts with limited resources or training, where practitioners experience competing demands, and can lead to variability in intervention effectiveness and usefulness.^26,27^ The challenges of implementing these interventions in Honduras during the COVID-19 pandemic elevated the importance of engaging a multi-disciplinary team in participatory intervention and implementation strategy design.^28^ Strategies focused on fidelity were prioritized to ensure the interventions reached their target audience as intended.^29^ This analysis describes and evaluates the fidelity of remote patient monitoring and self-administered pulse oximetry, focusing on implementation strategies, delivery quality, and participant responsiveness. In the future, remote patient monitoring and related technologies may become vital tools for expanding healthcare access, both during emergencies and in routine care. Findings from this study will help improve understanding of key factors that support implementation and inform recommendations for future interventions.

## Materials and methods

We conducted a retrospective analysis of the implementation fidelity of a block-randomized trial assessing phone-based remote patient monitoring and self-administered pulse oximetry for COVID-19 patients at high risk for severe disease – but who did not meet the criteria for hospitalization. Relying on the Conceptual Framework for Implementation Fidelity (CFIF), fidelity encompasses adherence (comprised of intervention content, coverage, frequency, and duration), while the relationship between the intervention and adherence may be moderated by intervention complexity, implementation strategies, delivery quality, and participant responsiveness.^26^ We conducted analyses for this cases study using existing data, including trial data and acceptability assessments; trial implementation notes; and compared those to expectations set out in the implementation handbook and standard operating procedures.

**Figure 1:**
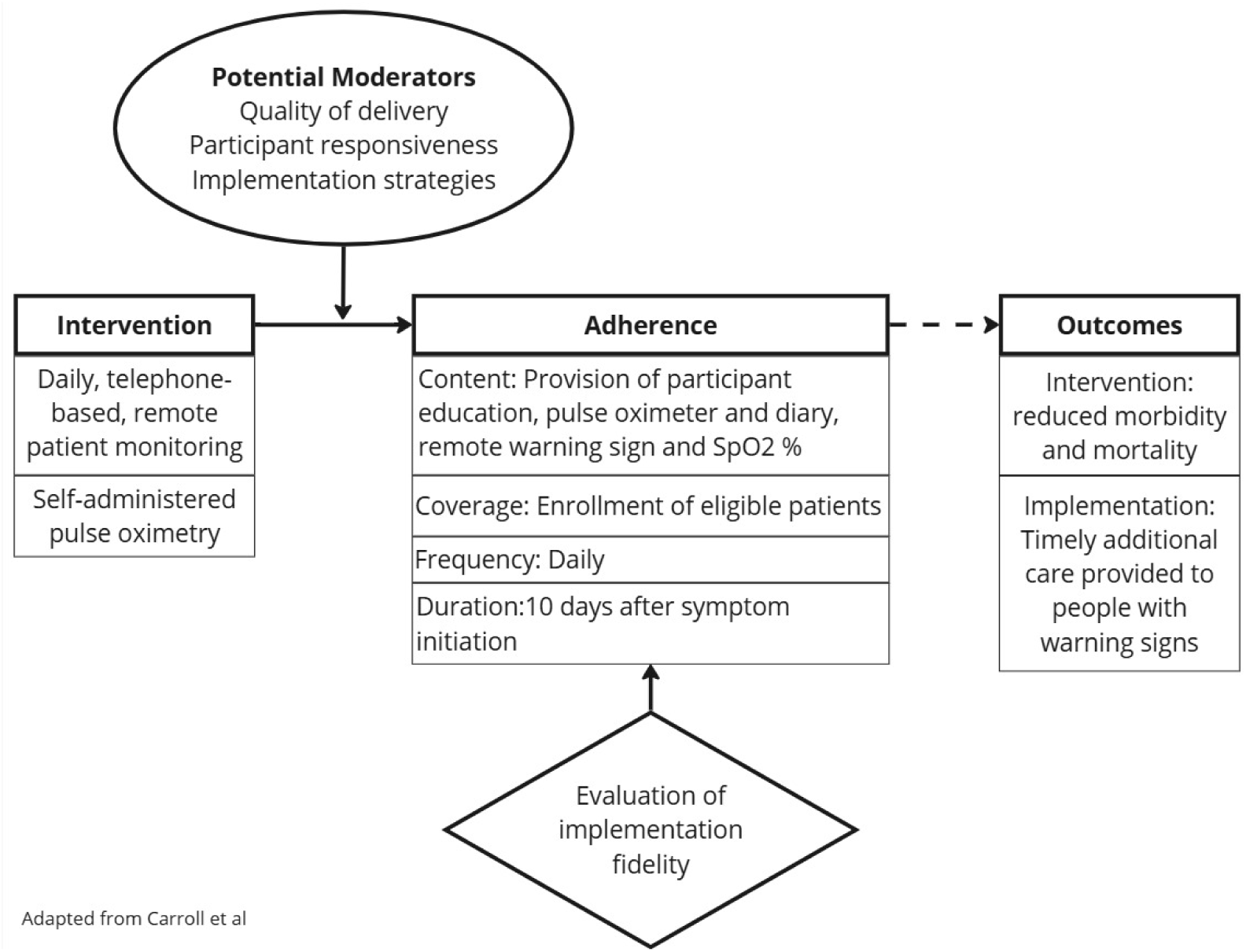
Adapted Conceptual Framework for Implementation Fidelity (CFIF)^26^. Sp0_2_ – Peripheral oxygen saturation, or blood oxygen

### INTERVENTION DESIGN

The interventions are described briefly to provide context. SESAL established COVID-19 triage centers in Tegucigalpa and Comayagüela, which were the primary public sector sites for COVID-19 diagnosis and care. Participants were SARS-CoV-2 positive via rapid antigen test, symptomatic, 60+ or 45-59 with CDC-defined high-risk conditions, and discharged home from a triage center. ^30^ Participants were enrolled by study-employed physicians at five COVID-19 triage centers managed by national authorities.

At enrollment, participants provided informed consent, were administered questionnaires, and received health education. Participants were randomized to receive daily, telephone-based remote monitoring, with or without a pulse oximeter. Those in the pulse oximetry group were given a pulse oximeter, usage instructions, and a diary for SpO_2_ readings. During monitoring calls, study-employed nurses assessed participant warning signs; if were identified, including SpO_2_ ≤ 94%, participants were referred for in-person evaluation. Monitoring was conducted for 10 days from symptom onset, unless participants reported ongoing fever, in which case monitoring continued until the participant was fever-free for 24 hours. Participants were asked to return pulse oximeters to triage centers for sterilization and re-use. The trial’s primary outcome was reduced mortality among participants using pulse oximeters, which is reported in an independent manuscript.

### FIDELITY EVALUATION

We examine the components of adherence using quantitative data collected during intervention implementation. (Table 1) Adherence was operationalized as relating to intervention delivery according to the study handbook.

**Table 1:** Intervention fidelity outcome measures.

| **Fidelity component** | **Fidelity outcome measures** |
| --- | --- |
| Adherence | Proportion of intervention **content** delivered as intended including: COVID-19 education, study supplies delivery at enrollment, screening for warning signs and low SpO<sub>2</sub> ($\leq 94\%$), and referral for additional care when identified<sup>1,3</sup> |
|  | **Coverage** of the target population, i.e. proportion of eligible patients enrolled<sup>1</sup> |
|  | **Frequency** of remote patient monitoring calls made by study staff versus frequency intended<sup>1</sup> |
|  | **Duration** of participant enrollment in the intervention versus enrollment intended<sup>1</sup> |

Potential moderators of the relationship between implementation and adherence include strategies, delivery quality, and participant responsiveness, which are interconnected. (Table 2) Implementation strategies aim to improve delivery quality and participant responsiveness, while delivery quality may affect responsiveness. Participant responsiveness was measured at each stage of the intervention where participants could choose to disengage.

**Table 2:**
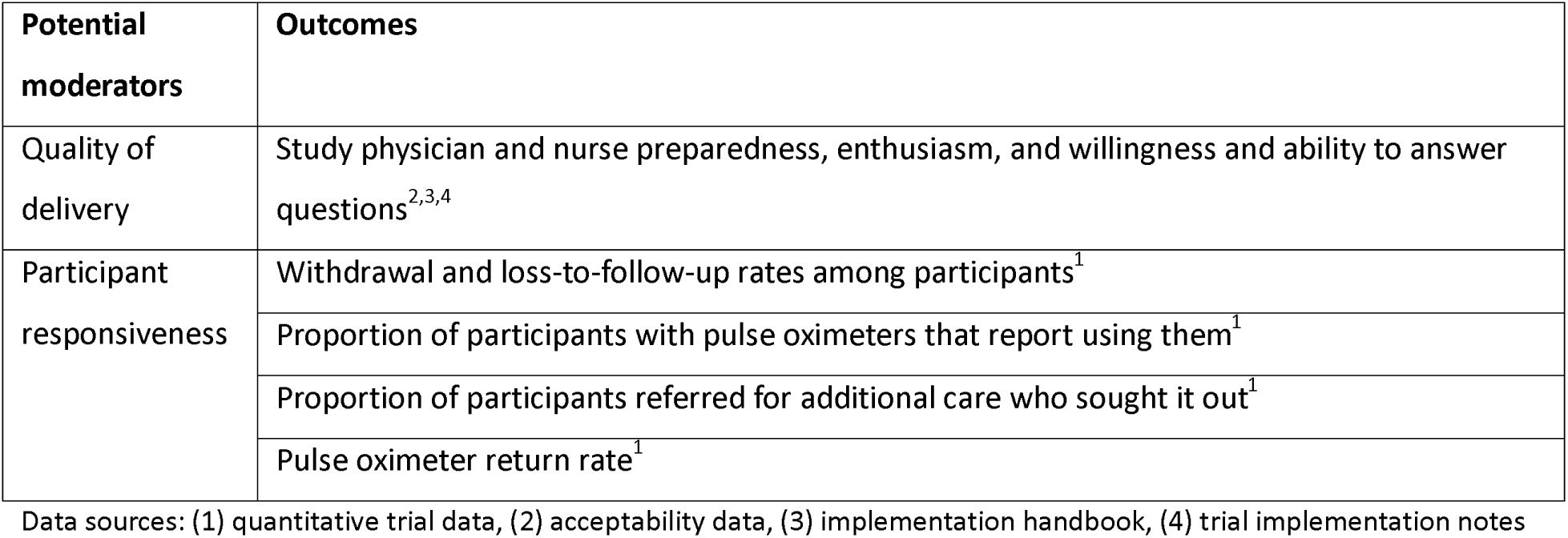
Outcomes of quality of delivery and participant responsiveness as potential moderators of the relationship between the intervention and adherence:

Implementation strategies were developed to improve adherence, delivery quality, and participant responsiveness. (Table 3) Many strategies targeted multiple areas of fidelity simultaneously. For example, piloting questionnaires with the target population helped ensure questions were easy to ask, supporting adherence, and easy to answer, promoting responsiveness and continued participation. Additionally, questionnaire piloting aimed to improve delivery quality by ensuring study staff had practice and received feedback from supervisors.

**Table 3:**
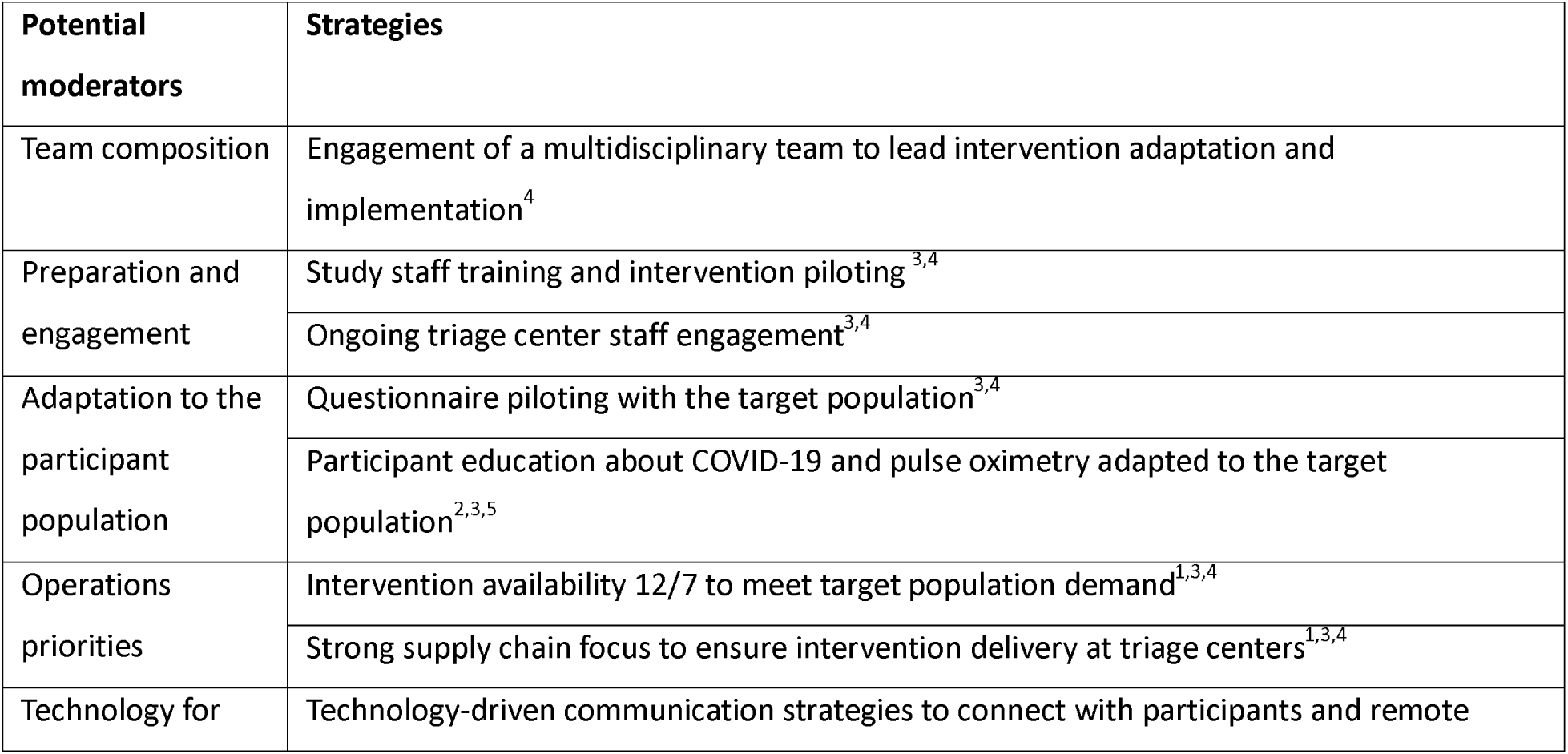
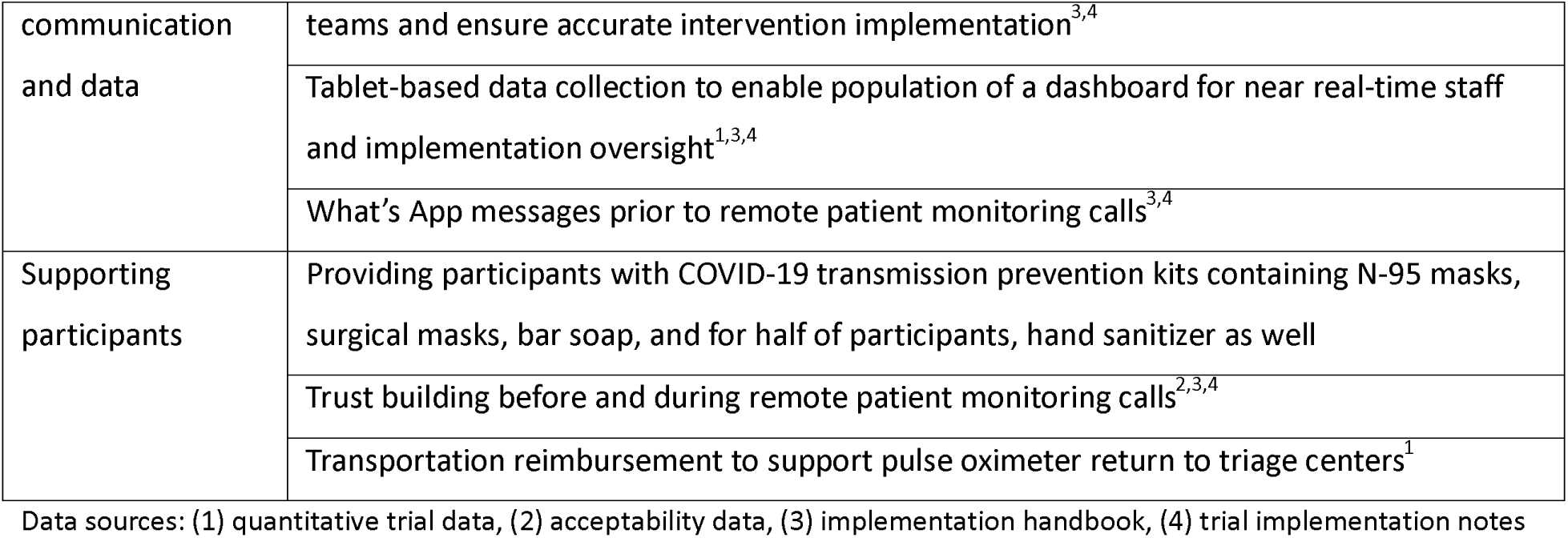
Implementation strategies employed to enhance quality of delivery and participant responsiveness.

### ANALYSIS

Quantitative analysis of acceptability and trial data involved the calculation of descriptive statistics, reported as frequencies and percentages. All analyses were conducted using STATA (version 17.0).^31^ Qualitative data were analyzed using deductive framework analysis, with space for inductive code generation.^32^ A document review was conducted on the implementation handbook, and implementation notes taken by study staff, focusing on fidelity analysis. The review assessed the alignment between the handbook and actual practices, as reflected in study data and implementation notes.

### ETHICAL APPROVAL

Institutional Review Board (IRB) review was received from the Massachusetts General Brigham (2021P001143) and the Autonomous University of Honduras (00003070). Secondary data analysis was approved by the Johns Hopkins Bloomberg School of Public Health (24586).

## Results

Implementation fidelity results related to adherence, quality of delivery, participant responsiveness, and implementation strategies are presented below.

### ADHERENCE

Assessment of adherence outcomes included: coverage, content, frequency, and duration (Table 1). The intervention achieved 97.9% coverage, enrolling 1,821 out of 1,860 eligible patients. Among those not enrolled (n=39), 19 did not qualify, 19 declined participation, and one was enrolled twice.

Intervention content was delivered consistently. All 1,821 patients received COVID-19 education and appropriate study supplies at enrollment, with no reported stockouts. Screening for warning signs and low SpO_2_ (≤94%)—where applicable—was conducted during each remote monitoring call. All participants reporting an SpO_2_ below ≤94% during a monitoring call (n=43) were referred. Of the 116 remote monitoring calls with reported warning signs, healthcare staff failed to refer participants on eight occasions (6.9%), despite the protocol mandating universal referral. Reasons participants were not referred included: one participant seeking care outside the triage system, one sought additional care the day prior, and another who reported the warning sign chronically, the remainder did not provide explanations. In response to non-referrals, staff received additional training to emphasize mandatory referrals, and form prompts and constraints were added to ensure compliance.

Frequency of remote patient monitoring calls aligned with expectations, each participant was called daily, and if not reached, called up to two more times that day. 12,602 (97.7%) daily monitoring calls were completed; 2.3% (n=295) were not answered. Seventy-three calls were made to people who could not be contacted and later classified as lost to follow-up, defined as not answering the monitoring call on three consecutive days. Due to an implementation error, two enrolled participants were called 10 times each without being reached; they were subsequently classified as lost to follow-up. Expected intervention enrollment duration was 10 days post-onset of COVID-19 symptoms – enrollment occurred a mean of 2.9 days (SD 1.5) after symptom onset and participation duration mean was 7.4 days (SD 1.4).

## Moderating factors

### QUALITY OF DELIVERY

Quality of delivery was assessed through staff supervision and participant feedback. Supervision demonstrated that staff adapted their educational and communication approaches based on participants’ reading and health literacy levels, and understanding of COVID-19. Findings were corroborated by participant responses to acceptability questions, with 99.7% (n=1,754) of respondents expressing satisfaction with the intervention, and 94.3% (n=1,667) willing to participate again if they contracted COVID-19. A small portion, 0.6% (n=11), were unsure, while 5.0% (n=89) indicated they would not participate again. Detailed intervention acceptability results can be found in an independent manuscript.

Senior study staff observed study physicians and nurses consistently using kind and empathetic communication techniques, such as greeting patients, speaking slowly and clearly, and answering participant questions, which was emphasized during training. They dedicated significant time to supporting participants and incorporating family members into education and monitoring, when appropriate.

### PARTICIPANT RESPONSIVENESS

Participant responsiveness was assessed through study withdrawal and loss to follow-up, pulse oximeter use, responsiveness to referrals for additional care, and pulse oximeter recapture rate.

Of 1,821 participants enrolled, 0.1% (n=2) withdrew, and 2.0% (n=37) were lost to follow-up. Those who withdrew described phone calls as bothersome. Participant engagement was evident in the high rates of successful remote patient monitoring calls, described earlier. 97.8% (n=877) of participants who received a pulse oximeter reported using it as directed during enrollment.

Participant responsiveness to referrals for additional care after warning signs or an SpO_2_≤ 94% was lower than expected. Overall, 100 participants were referred for follow-up during 123 calls where warning signs were reported; participants were re-referred on subsequent calls if symptoms persisted. Of these, 52 reported seeking care or the outcome of their referral remained unknown. During chart review to determine final outcomes, study staff followed up with participants when no chart or other evidence of return for care could be found. During follow-up calls, 24 participants revised their initial responses, indicating that they had not sought additional care. Ultimately, 28% (n=28) of participants sought care after being referred, with 27 attending SESAL or IHSS health facilities and one attending a private health facility.

Pulse oximeter recapture aimed to enhance the intervention’s cost-effectiveness. Participants were asked to return devices to be sterilized and redistributed. To encourage returns, participants were offered ∼$10 USD to cover transportation expenses. Of the 500 pulse oximeters purchased, 283 were not returned after discharge of 897 participants in the pulse oximeter arm, a recapture rate of 68.5%.

### IMPLEMENTATION STRATEGIES

#### Team composition

The multi-disciplinary study team consisted of public health practitioners, medical doctors, epidemiologists, program managers, and a microbiologist. The team worked closely with key stakeholders from national, departmental, and city-level SESAL offices, as well as COVID-19 triage centers. In-person and virtual stakeholder meetings were held regularly to ensure consistent communication and coordination.

#### Preparation and engagement

Study staff underwent five days of training and intervention piloting, led by SESAL and study staff. Training included problem-solving exercises and role-playing participant interactions to refine communication techniques and ensure high intervention quality. Job aids outlining intervention participation criteria were posted in exam rooms to support triage physicians, and paper invitations were given to patients to encourage them to visit the study enrollment site within the triage center after referral by a triage center physician. Only 1.0% (n=19) of referrals were erroneous.

#### Adaptation to the participant population

Piloting the study’s questionnaires with potential participants helped ensure questions were easy to understand. One notable issue, was a question about whether participants felt their health had returned to its pre-COVID baseline, which led to confusion, with participants often reporting that they felt better after versus before their COVID-19 illness, despite reporting persistent symptoms. As a result, this question was excluded from analysis.

During enrollment, study physicians delivered education about COVID-19 transmission, warning signs, and pulse oximeter use. Staff engaged family members who accompanied participants to reinforce study instructions at home. A printed SpO_2_ diary with visual and written instructions, along with an in-person demonstration, were provided to enable thrice-daily readings, recording, and reporting them during remote monitoring calls.

#### Operations priorities

A staggered rollout approach was used for implementation. On the first day, the study team and SESAL collaborators, attended a single triage center to practice skills, observe operations, and provide feedback to ensure consistent and quality implementation. Additional study sites were added over one week. Once fully operational, study physicians were stationed at COVID-19 screening sites 12 hours a day, 7 days a week. Two triage centers closed during the study, and one enrollment site was added at the Social Security Hospital. Enrollment schedules were adjusted when COVID-19 rates declined between waves, reducing staffing and expenses to allow intervention extension.

Ensuring daily supplies for enrollment sites was a complex and essential task. During implementation every participant received the appropriate supplies at the time they were enrolled, thanks to a study coordinator who managed a central commodities store, inventory at remote enrollment sites, and could quickly provide additional supplies when needed, supported by a dedicated study vehicle and driver.

#### Technology for communication and data

Phone-based monitoring was effective throughout the trial. Concerns about phone ownership among participants were allayed due to the urban setting and feedback from study staff. Among participants, 92.9% (n=1,692) owned a phone, 6.8% (n=124) borrowed one from someone in their household, and 0.3% (n=5) borrowed from someone outside their household. Of 19 qualified patients who declined enrollment, one person reported it was due to lack of telephone access. Phone ownership declined significantly with age—participants aged 85 and older were 52 times more likely to report using someone else’s phone to participate than participants 45 – 50 years old (OR 52.99 [CI 20.81 – 134.96]). Age was not associated with likelihood of enrollment.

WhatsApp, which is highly accessible and included with pre-paid phone credits in Honduras, was an effective communication tool, supporting smooth operations and used in participant communication. Daily reminders for staff addressed enrollment logistics, staffing and supervision, changes at triage centers, supply management, and clarification of enrollment criteria. Initially, when participants were referred for additional care, triage center staff had no way to know the patient was part of the stud, or which warning sign had triggered the referral. In response, digital referral slips were introduced, identifying the participant as part of the study and listing the referral reason. These digital slips were sent to participants via WhatsApp.

Data were collected using Kobo Collect on tablets and uploaded daily to a secure cloud-based server via secure Wi-Fi. Automated spreadsheets tracked participants and daily call lists, while a Power BI dashboard facilitated supervision and information sharing among the study team, implementing team, and SESAL staff. Regular data reviews helped monitor protocol adherence, including enrollment, participant inclusion criteria, delivery of intervention components, enrollment days, and referrals after warning signs were reported. As protocol deviations were identified, pop-up warnings and constraints were added to electronic forms to reinforce compliance.

#### Participant support

The provision of COVID-19 transmission prevention kits, which contained N-95 and surgical masks, bar soap, and alcohol-based hand rub (distributed to 50% of participants), may have enhanced study enrollment. The timing of the study coincided with a period of decreased COVID-19 concern, suggesting that participants who actively sought diagnosis were health-conscious and thus may have been particularly receptive to receiving preventive supplies as a study incentive.

Establishing participant trust was essential to encourage continued engagement and maintain quality delivery. This process began by adhering to familiar Honduran greeting styles and communication patterns, allowing sufficient time for participant interaction to ensure understanding and a trusting relationship. As mentioned previously, sending WhatsApp messages to study participants prior to the monitoring phone calls further strengthened trust by reassuring participants of the legitimacy of the calls. Ensuring that all aspects of the intervention were delivered as promised—from providing supplies during enrollment to initiating daily monitoring calls at the participant’s preferred time and issuing referral slips—was key to building confidence in the study and fostering engagement.

At enrollment, participants were asked to return pulse oximeters after participation. Study nurses reminded participants on their final day of enrollment and followed up through additional calls, if time allowed. Messaging encouraged participants to return pulse oximeters to aid the study and to benefit their fellow Hondurans. Upon return, participants received a phone credit valued at approximately $10 USD to cover transportation costs.

## Adaptation in line with emerging evidence

From study conceptualization in 2021 to conclusion January 2023, key changes occurred. Shortly after study inception, the CDC updated the pre-existing conditions linked to COVID-19 risk (when five participants were enrolled); thereafter hypertension was added as a qualifying condition. Additionally, the minimum enrollment age was lowered from 50 to 45 years for those with comorbidities, and from 65 to 60 years for those without. After emerging evidence found that pulse oximeters overestimate SpO_2_levels by 1.1-1.2% in people with darker skin, the study team emphasized mandatory referral after warning signs reports, including if SpO_2_ was > 94%.^33^

## Harms and unintended effects

No unintended harms from study participation or interventions were identified. Unexpected positive impacts on mental and emotional health during COVID-19 illness and isolation were widely reported by participants to nurses and study staff when returning equipment and completing questionnaires. Healthcare providers noted these positive effects during interviews, with some believing this was a study objective. SESAL providers reported the intervention may have reduced triage center burden by decreasing unnecessary care visits, though corroborating data are unavailable.

## Discussion

This analysis demonstrated high implementation fidelity across key metrics, achieving 97.9% coverage, consistent delivery of COVID-19 education and supplies, 97.7% completion of prescribed remote monitoring calls, and 97.8% of participants using pulse oximeters. Despite 28% of referred participants seeking additional care, 99.7% were satisfied with the intervention. Implementation strategies, including population-tailored education materials and WhatsApp communication, contributed to fidelity. Adaptations such as staff training and evidence-based modifications maintained intervention quality. The pulse oximeter return rates was 68.5%. Qualitative feedback indicated unexpected emotional and mental health benefits during COVID-19 isolation.

Although implementation strategy testing was not the primary focus of the parent trial, we identified high fidelity, enrollment rates, and patient responsiveness. Concerns about remote patient monitoring in LMIC typically focus on reliable electricity, mobile networks, technology access and skills, especially among the marginalized groups, and cost.^8,34–42^. However, our findings contrast these concerns, as participants successfully used telephones, pulse oximeters, and reported warning signs. A seven-country systematic review of remote monitoring during COVID-19 found patient engagement was a frequent fidelity gap and a similar UK program had very low intervention enrollment (1.17%) among eligible patients, undermining results.^34,43^ The high participant responsiveness seen here could be context- or population-specific, such as greater willingness to receive calls due to isolation or the older population, warranting further investigation with other populations and settings.

Quality of delivery, described as an “obvious potential moderator” of the relationship between an intervention and fidelity, likely impacted patient responsiveness.^26^ Responsiveness was crucial at each intervention step, from enrollment, to phone calls, describing warning signs, using pulse oximeters, acting on referrals, and returning pulse oximeters. Evidence suggests a reciprocal relationship where patient engagement inspires providers to deliver quality care by encouraging collaborative decision-making and communication.^44,45^ Minor adjustments based on participant feedback, like sending text messages before calls and engaging in culturally appropriate conversation, may have positively impacted responsiveness without reducing fidelity.^45^ These approaches built trust, which has been shown to promote health system engagement and improve clinical outcomes.^46–48^

Among similar studies, we identified one trial that published patient and provider perspectives of a text message-based approach implemented in Pennsylvania, USA,.^49^ Key challenges included technology use, especially among the elderly, low literacy, non-English languages, and unclear guidance on seeking care. Recommendations emphasized including phone calls to allow provider assessment, adaptation to diverse populations including, and enhancing accessibility through implementation strategies. These reflections reinforce the importance of engaging a multidisciplinary team to develop implementation strategies with input from the target population prior to implementation.

We did not identify other similar studies that reported adherence to referral recommendations or strategies to facilitate return. In our intervention, low adherence to referral recommendation was the only step with lower-than-expected responsiveness. In hindsight, few implementation strategies were employed beyond nurse encouragement and digital referral slips. Understanding why participants more readily returned pulse oximeters than sought care after warning signs could offer insight. Possible explanations offered during healthcare provider interviews included improved health status to travel, avoiding wait times see a healthcare provider, or receiving travel reimbursement, but a clear understanding of barriers and approaches merit further investigation.

### Strengths and limitations

This study’s strengths include the participatory design, multi-disciplinary team input, and the iterative and adaptive process. Discussions with key stakeholders and study staff led to substantive changes to intervention and implementation design. While the use of human-centered design was not an explicit goal, placing participants at the center, speaking with members of the target population, anticipating challenges they might face, and prioritizing participation accessible participation reflect that approach.

However, there are limitations. First, the parent trial was not designed as a fidelity study, so results are observational. Second, knowledge about COVID-19, silent hypoxia, and pulse oximetry evolved during the trial, creating challenges in balancing new insights with maintaining protocol fidelity. Third, quality of delivery was assessed subjectively, without consistent application of a validated framework, which could introduce recall and familiarity bias. Finally, no data are available to verify perceived reduction in health facility burden and perceived impact on participant mental and emotional health.

Future research on remote patient monitoring and self-administered pulse oximetry should prioritize hybrid effectiveness-implementation trials that test both interventions and implementation strategies simultaneously. Given the disparities in implementing success across settings, evaluating the identified strategies could provide evidence about their influence on outcomes. Before deploying a self-guided approaches for follow-up care, researchers should engage with target populations to understand physical, social, and economic barriers, then develop and measure strategies to address those barriers to promote access and equity.

This approach has broad applications for healthcare systems in Honduras and beyond, particularly during health emergencies. Our success with remote monitoring in a setting previously unfamiliar with telehealth demonstrates the potential for scaling telehealth solutions in both emergency and primary care contexts. As the Pan American Health Organization prioritizes digital health transformation, we must focus on how telehealth can improve access for marginalized populations and leveraged during outbreaks and other public health emergencies.^50^ However, as demonstrated, accounting for technology access and incorporating the support available within multigenerational households and community networks can contribute to feasibility. These interventions brought healthcare closer to patients and provided guidance on seeking care during acute respiratory illness. Such remote monitoring can help overcome access barriers during emergencies and among marginalized groups facing barriers related to limited mobility, transportation costs, or mistrust of traditional healthcare settings.

## Conclusions

This investigation discusses the introduction of remote patient monitoring and self-administered pulse oximetry in Tegucigalpa and Comayagüela, Honduras, during the COVID-19 health emergency—marking the first use of these approaches in these cities. By leveraging local expertise and public health insights, context-appropriate implementation strategies were developed. The successful implementation of these interventions demonstrates the potential for expanding them in other LMICs to address ongoing healthcare access challenges. Further research into the generalizability and scalability of these methods, both as enhancements to existing care pathways and as tools for pandemics and other health emergencies, could improve healthcare access and support digital health transformation.

## Data Availability

Data belong to the Secretariat of Health of Honduras and will be shared upon reasonable request.

## Acknowledgements

The authors would like to thank the residents of Tegucigalpa and Comayagüela, Honduras who consented and participated in the study. We thank the nurses and doctors employed by the trial who worked tirelessly to implement these interventions. We thank the city, regional, and national members of the Honduras Secretary of Health and the Hospital Institute for Social Security.

## Author contribution statement

Kathryn W. Roberts: Conceptualization, Data curation, Formal analysis, Investigation, Methodology, Supervision, Validation, Visualization, Writing – original draft preparation; Berta Alvarez: Conceptualization, Investigation, Methodology, Project administration, Supervision, Writing – review and editing; Michael de St. Aubin: Data curation, Investigation, Project administration, Software, Writing – review and editing; Omar Diaz: Conceptualization, Investigation, Methodology, Project administration, Supervision, Writing – review and editing; Salomé Garnier: Investigation, Software, Writing – review and editing; Rachel See: Methodology, Supervision, Project administration, Writing – review and editing; Saul Cruz: Investigation, Supervision, Writing – review and editing; Lorenzo Pavon: Investigation, Supervision, Writing – review and editing; Anthony So: Methodology, Supervision, Writing – review and editing; Ligia Paina: Methodology, Supervision, Writing – review and editing; Tara Kirk Sell: Methodology, Supervision, Writing – review and editing; Shiony Midence: Investigation, Supervision, Writing – review and editing; Angela Ochoa: Investigation, Supervision, Writing – review and editing; Homer Mejía Santos: Investigation, Supervision, Writing – review and editing; Jonatán Ochoa: Investigation, Supervision, Writing – review and editing; Sogeiry Solis: Investigation, Methodology, Project administration, Supervision, Writing – review and editing; Devan Dumas: Investigation, Methodology, Project administration, Supervision, Writing – review and editing; Margaret Baldwin: Investigation, Project administration, Writing – review and editing; C. Daniel Schnorr: Conceptualization, Writing – review and editing; Alcides Martinez: Conceptualization, Supervision, Writing – review and editing; Avi Hakim: Conceptualization, Methodology, Project administration, Supervision, Writing – review and editing; Eric Nilles: Conceptualization, Funding acquisition, Investigation, Methodology, Project administration, Supervision, Writing – review and editing.

## Funding statement

Funded by the U.S. Centers for Disease Control and Prevention, Cooperative Agreement Number U01GH002238 via Centers for Disease Control and Prevention, Central America Country Office, Guatemala; Reducing the morbidity and mortality due to acute febrile illnesses in Central America and the Dominican Republic. The trial was sponsored by the US CDC, which consulted on study design, analysis, and interpretation. All decisions were taken by the primary investigator.

Roche SARS-CoV-2 Rapid Antigen Tests were provided through an Investigator Initiated Study with Roche Diagnostics, Ltd, which played no role in study design, data collection, analysis, interpretation, writing of the report, or publication.

## Ethics statement

This activity was determined to meet the definition of research [45 CFR 46.102(l)] involving human subjects [45 CFR 46.102 (e)(1)] and Institutional Review Board (IRB) review and was approved by Massachusetts General Brigham Institutional Review Board (2021P001143), the Autonomous University of Honduras (00003070), and SESAL. Secondary data analysis was approved by the Johns Hopkins Bloomberg School of Public Health Institutional Review Board (Approval No. 24586). The protocol was reviewed by the U.S. Centers for Disease Control and Prevention and determined to be research but the CDC was not engaged.

## Data access statement

The data analyzed to prepare this manuscript are housed at the Secretariat of Health of Honduras and the Harvard Humanitarian Initiative and are available upon reasonable request to the corresponding author.

## Centers for Disease Control and Prevention (CDC) Disclaimer

The findings and conclusions in this report are those of the author(s) and do not necessarily represent the official position of the U.S. Centers for Disease Control and Prevention/the Agency for Toxic Substances and Disease Registry.

